# Abnormal Upregulation of Cardiovascular Disease Biomarker PLA2G7 Induced by Proinflammatory Macrophages in COVID-19 patients

**DOI:** 10.1101/2020.08.16.20175505

**Authors:** Yang Li, Yongzhong Jiang, Yi Zhang, Naizhe Li, Qiangling Yin, Linlin Liu, Xin Lv, Yan Liu, Aqian Li, Bin Fang, Jiajia Li, Hengping Ye, Gang Yang, Xiaoxian Cui, Yang Liu, Yuanyuan Qu, Chuan Li, Jiandong Li, Dexin Li, Shiwen Wang, Zhongtao Gai, Faxian Zhan, Mifang Liang

## Abstract

**BACKGROUND:** Coronavirus disease 2019 (COVID-19) triggers distinct patterns of pneumonia progression with multiorgan disease, calling for cell- and/or tissue-type specific host injury markers.

**METHODS:** An integrated hypothesis-free single biomarker analysis framework was performed on nasal swabs (n = 484) from patients with COVID-19 in GSE152075. The origin of candidate biomarker was assessed in single-cell RNA data (GSE145926). The candidate biomarker was validated in a cross-sectional cohort (n = 564) at both nucleotide and protein levels.

**RESULTS:** Phospholipase A2 group VII (PLA2G7) was identified as a candidate biomarker in COVID-19. PLA2G7 was predominantly expressed by proinflammatory macrophages in lungs emerging with progression of COVID-19. In the validation stage, PLA2G7 was found in patients with COVID-19 and pneumonia, especially in severe pneumonia, rather than patients suffered mild H1N1 influenza infection. Up to 100% positive rates of PLA2G7 were positively correlated with not only viral loads in patients with COVID-19 but also severity of pneumonia in non-COVID-19 patients. Although Ct values of PLA2G7 in severe pneumonia was significantly lower than that in moderate pneumonia (*P* = 7.2e-11), no differences were observed in moderate pneumonia with COVID-19 between severe pneumonia without COVID-19 (*P* = 0.81). Serum protein levels of PLA2G7, also known as lipoprotein-associated phospholipase A2 (Lp-PLA_2_), were further found to be elevated and beyond the upper limit of normal in patients with COVID-19, especially among the re-positive patients.

**CONCLUSIONS:** We firstly identified and validated PLA2G7, a biomarker for cardiovascular diseases (CVDs), was abnormally enhanced in COVID-19 patients at both nucleotide and protein aspects. These findings provided indications into the prevalence of cardiovascular involvements seen in COVID-19 patients. PLA2G7 could be a hallmark of COVID-19 for monitoring disease progress and therapeutic response.

**FUNDING:** This study was supported by grants from China Mega-Projects for Infectious Disease (2018ZX10711001), National Natural Science Foundation of China (82041023).

## Introduction

Severe acute respiratory syndrome coronavirus 2 (SARS-CoV-2) is a novel enveloped RNA betacoronavirus that emerged in December 2019 in Wuhan, China, and is the causative etiology of coronavirus disease 2019 (COVID-19)(1, 2). Typical clinical presentation of COVID-19 was a lung involvement, as evidenced by image tests, with fever, cough and dyspnea(3, 4). Severe cases often developed acute respiratory distress syndrome (ARDS) or even death(3, 4). The risk factors for increased disease severity in patients with COVID-19 has been reported as older age (e.g., over 50 years old) and the presence of comorbidities, including hypertension, diabetes mellitus, cardiovascular disease etc.(5, 6). However, it rapidly became obvious that severe COVID-19 can also occur in younger patients with no pre-existing comorbidities(7).

On-going data characterizing immunological features in patients with COVID-19 were starting to emerge. Host response to SARS-CoV-2 infection was distinct in comparison with other highly pathogenic coronaviruses and common respiratory viruses in cell lines(8). Notably, reduced type I interferon activities were observed in both in vitro(8) and in vivo(9) data. Delayed production of type I interferon resulted in boosted cytopathic effects (CPE) and increased sensing of SARS-COV-2 threats promoted the enhanced release of monocyte chemoattractants, such as CCL2(10), which contributed an influx of monocytes into lungs(7). Autopsy studies have shown diffuse thickening of the alveolar wall with mononuclear cells and macrophages infiltrating airspaces(11). However, it is still mysterious why there were various complications of COVID-19, including impaired function of the organs, especially heart(7). Among 1,216 hospitalized COVID-19 patients across 69 countries who did not have pre-existing heart complications, almost half showed scan abnormalities that resemble the early stages of heart failure(12).

Hypothesis-free biomarker studies could allow researchers to gain in depth insights into patho-mechanisms underlying COVID-19. Several attempts have been made. Expression of monocyte CD169 (mCD169), also known as sialic acid binding Ig like lectin 1 (SIGLEC1), has been suggested as a biomarker in the early diagnosis of COVID-19(13). But expression of SIGLEC1 could be enhanced by other viruses(14). On the other hand, protein-level biomarkers that could predict the severity of COVID-19 patients were identified(15, 16). In particular, the level of lactate dehydrogenase (LDH) in the blood which was used to monitor the tissue damage was highly indicative of COVID-19 mortality with area under curve (AUC) > 0.9(15). These results suggested the development of additional cell- and/or tissue-type specific host injury markers were needed(17). Here, we started to employ the previously established integrated hypothesis-free single biomarker analysis framework(18) on bulk transcriptomic data (n = 484) from nasal swabs retrieved from COVID-19 patients and controls. To further understand the contribution to COVID-19 of this gene, single cell RNA (scRNA) data from bronchoalveolar lavage fluids (BALFs) of moderate and severe COVID-19 patients were also explored. Thereafter, more than 500 clinical samples, including nasal swabs and serum samples, were collected to validate our findings at nucleotide and protein levels.

## Results

### In silico discovery of PLA2G7 as the biomarker of SARS-COV-2 infection

The clinical records in GSE152075 labelled as ‘not collected’ were removed. Batch-corrected count matrix was normalized by varianceStabilizingTransformation (vst) function in R package DEseq2(19). Following cluster analysis on the preprocessed expression matrix to remove outliers, in a total of 428 samples out of 484 were remaining (Supplementary Figure S1). As the scale-free topology fit index failed to reach values above 0.85 (Default Threshold), the soft-thresholding power of 12 was selected(20) (Supplementary Figure S2). In a total number of 8 modules were identified and constructed by WGCNA(21) analysis (Figure 1A; Supplementary Figure S3). To identify the SARS-COV-2 infection-related module, the Pearson correlation analysis, which involved calculating the Student asymptotic *P*-values for the correlations, between the module eigengenes (MEs) of each module and clinical traits was performed (Figure 1B). The red module was the module most relevant to SARS-COV-2 infection. Genes in red module (n = 60) were listed in Supplementary Table 1. Importantly, the disease ontology (DO)(22) analysis on these genes suggested not only upper respiratory tract disease, but also heart disease related terms (e.g., congestive heart failure and myocardial infarction) (Figure 1C). In addition, the network connections among the most connected genes in the red module was displayed through Cytoscape(23) (Figure 1D).

**Figure 1.**
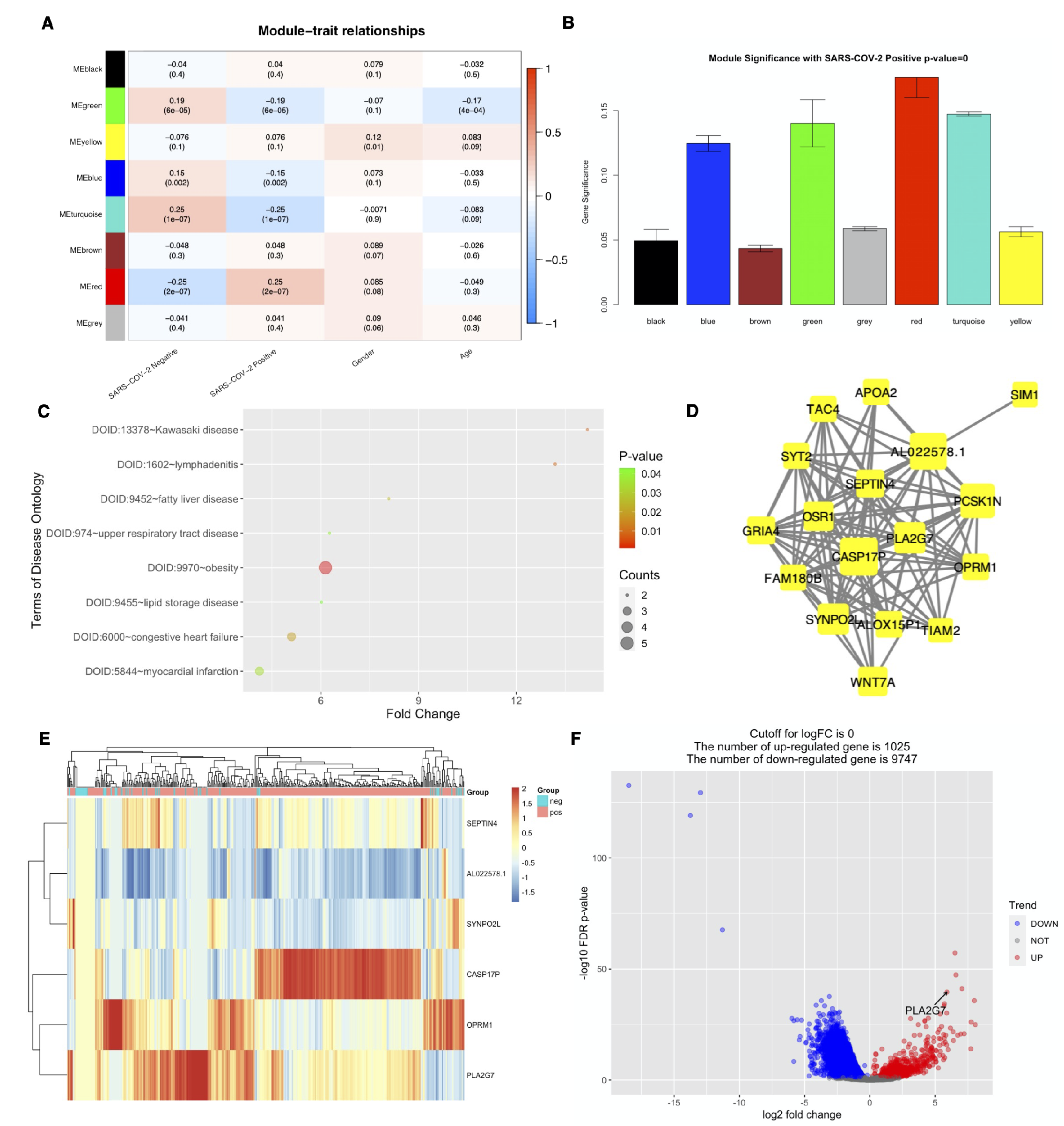
In silico discovery of PLA2G7 as the biomarker of SARS-COV-2 infection. (**A**) Heatmap of the correlation between module eigengenes and the clinical traits recorded in GSE152075. (**B**) Distribution of average gene significance and errors in the modules associated with positivity of SARS-COV-2. (**C)** Disease Ontology (DO) enrichment results on genes in red module. (**D)** Visualization of the network connections among the most connected genes in the red module. The size of circles was equal to the log2 fold change. (**E**) Heatmap based on unsupervised clustering of the selected 6 genes after scaling. Each row represented one gene; each column represents one patient. Expression intensity is indicated by color. (**F**) Volcano plot of differentially expressed genes (DEGs) for SARS-COV-2 infection contrasting positive cases with negative ones in GSE152075.

The genes in the red module were considered as candidate hub genes in SARS-COV-2 infection. In a total of 6 genes were selected based on a gene significance threshold of 0.8 and a module membership significance of 0.25 (Figure 1E; Supplementary Figure S4). Next, hub gene selection based on XGBoost(24) was carried out. First of all, all the samples were randomly assigned at a 7:3 ratio to a training set (299 samples) and a test set (129 samples). The python packages “XGBoost”(24) and “scikit-learn”(25) were used for classification. Second, to obtain the best XGBoost model parameter combination (eta, gamma, max_depth, Min_child_weight, Subsample, and n_ Colsample_bytree) with the highest classification accuracy, five-fold cross-validation and grid search were applied to the training set. Finally, the highest accuracy of classification was 0.876 which could be achieved through a single gene, phospholipase A2 group VII (PLA2G7) (Supplementary Figure S5). Moreover, the AUC score by R package “pROC”(26) in the training and test sets on this single gene was 0.850 and 0.820, respectively (Supplementary Figure S6). It should be noted that PLA2G7 was significantly upregulated in SARS-COV-2 positive group (logFC = 5.903, FDR *P*-value = 2.68e-40) (Figure 1F).

### PLA2G7 was induced by proinflammatory macrophages emerging along with progression of COVID-19

Clustering analysis on scRNA-seq data (GSE145926) showed 27 distinct clusters composed of macrophages, neutrophils, myeloid dendritic cells (mDCs), plasmacytoid dendritic cells (pDCs), natural killer (NK) cells, T cells, B cells, plasma cells and epithelial cells (Figure 2A, Supplementary Figure S7–8). The gene expression analysis showed PLA2G7 was expressed principally by macrophages (Figure 2A-B). To further understand the origin of PLA2G7, the 15 macrophages subclusters were re-integrated and re-clustered (Supplementary Figure S9). The expression patterns of PLA2G7, along with typical classification markers of macrophages (FCN1, SPP1 and FABP4)(27, 28), were then explored (Figure 2C). Thereafter, four heterogeneous subgroups of macrophages were identified, FCN1^hi^ group, FCN1^lo^SPP1^+^ group, SPP1^+^ group and FABP4^+^ group, those were consistent with previous study(27)(Supplementary Figure S10–11). It was worth of noting that the expression profile of PLA2G7 shared a close pattern to SPP1 (Figure 2D), which played an important role in idiopathic pulmonary fibrosis(28). Macrophages with FABP4, also known as the alveolar macrophages, were reduced with progression of COVID-19 while macrophages with FCN1 and SPP1, considered as the proinflammatory macrophages, were strikingly increased (Figure 2E). In addition, strong correlation between expression of PLA2G7 and progression of COVID-19 was discovered (r = 0.927, *P* = 1.4e-4) (Figure 2F). On the whole, PLA2G7 was predominantly expressed by these proinflammatory macrophages emerging along with progression of COVID-19.

**Figure 2.**
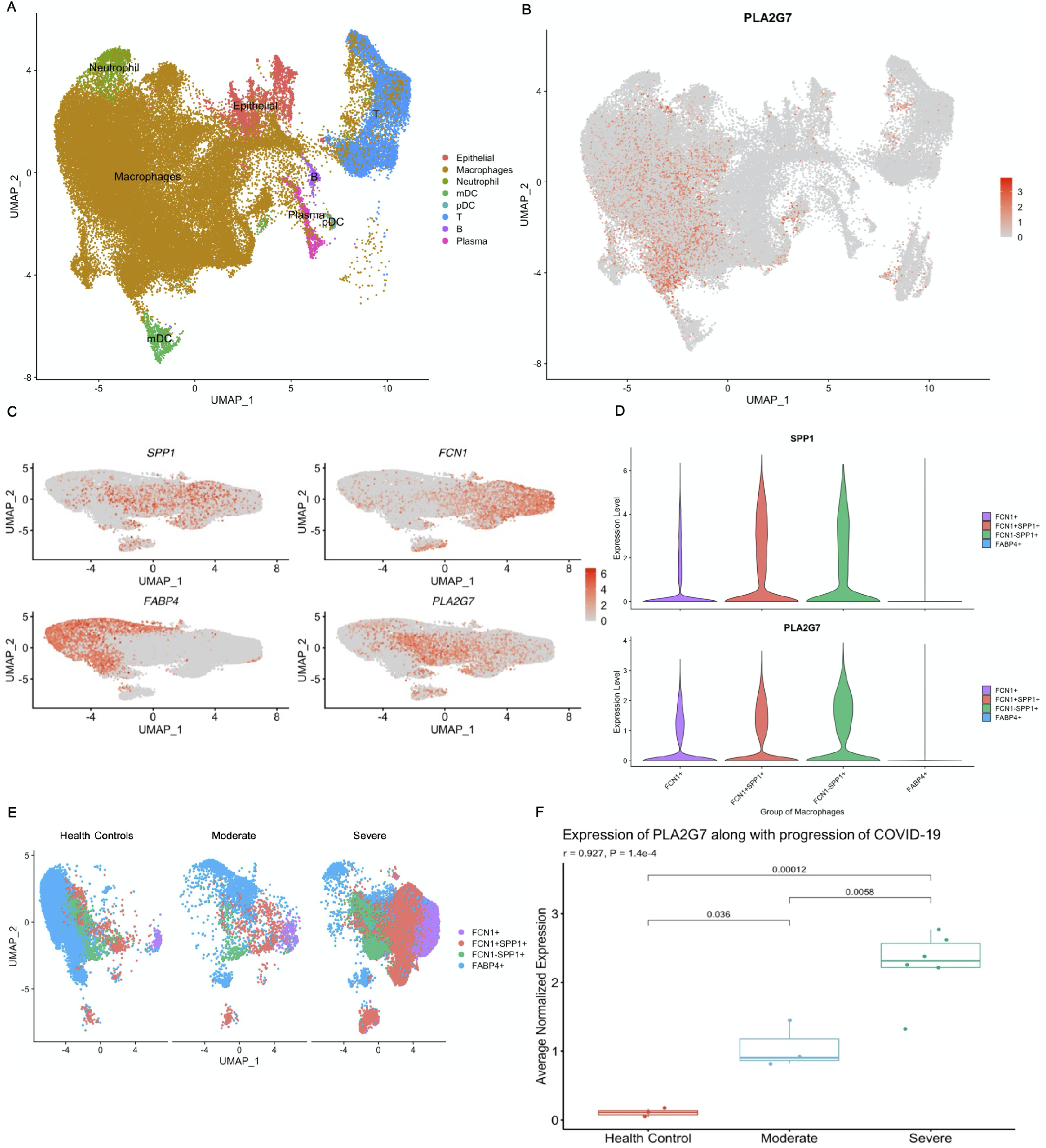
PLA2G7 was induced by proinflammatory macrophages emerging along with progression of COVID-19. **(A)** UMAP presentation of major cell types in bronchoalveolar lavage fluids (BALFs) from COVID-19 patients in GSE145926. **(B)** UMAP plot showing the expression of PLA2G7 across the immune cells. (**C**) UMAP maps showing the expression of PLA2G7, along with the markers (SPP1, FCN1 and FABP4) in re-integrated macrophages. **(D)** Expression of SPP1 and PLA2G7 by four macrophages groups. **(E)** UMAP projection of four macrophage groups among the health controls, moderate and severe COVID-19 patients. **(F)** Average normalized expression of PLA2G7 was correlated with progression of COVID-19. UMAP, Uniform Manifold Approximation and Projection.

### PLA2G7 was detected in a retrospective collection of nasal swabs from both COVID-19 and pneumonia patients

In a total of 447 nasal swabs were collected and subject to SARS-COV-2 targeted qRTPCR. The 447 nasal swabs were grouped as: Health controls (n = 200), COVID-19 group (n = 134), Influenza infection (n = 52), Severe pneumonia (n = 41) and Moderate pneumonia (n = 20). It should be noted the median (interquartile range [IQR]) of Ct of COVID-19 group in present study were relatively higher than that in public dataset GSE152075 (SARS-COV-2 Ct: 31.66 [28.62 – 34.00] vs. 21.31 [19.09 – 24.01]) (Figure 3A). To match the records in GSE152075, the COVID-19 group were divided into two groups with Ct value ≤ 25 and > 25. Thereafter, we next explored the relationship between positive perdition value (PPV) of PLA2G7 and Ct values of SARS-COV-2 in COVID-19 group. The PPV of PLA2G7 was correlated with Ct of SARS-COV-2 (r = –0.96, p = 3.3e-12) (Figure 3B), indicated that the PPV of PLA2G7 was positively correlated with SRAS-CoV-2 viral load. It was worth of noting more than 80% PLA2G7 positive rate were observed in COVID-19 patients with Ct of SARS-COV-2 ≤ 25 (Figure 3B-C), which showed comparable diagnostic performance in GSE152075 as shown in Figure 1E.

**Figure 3.**
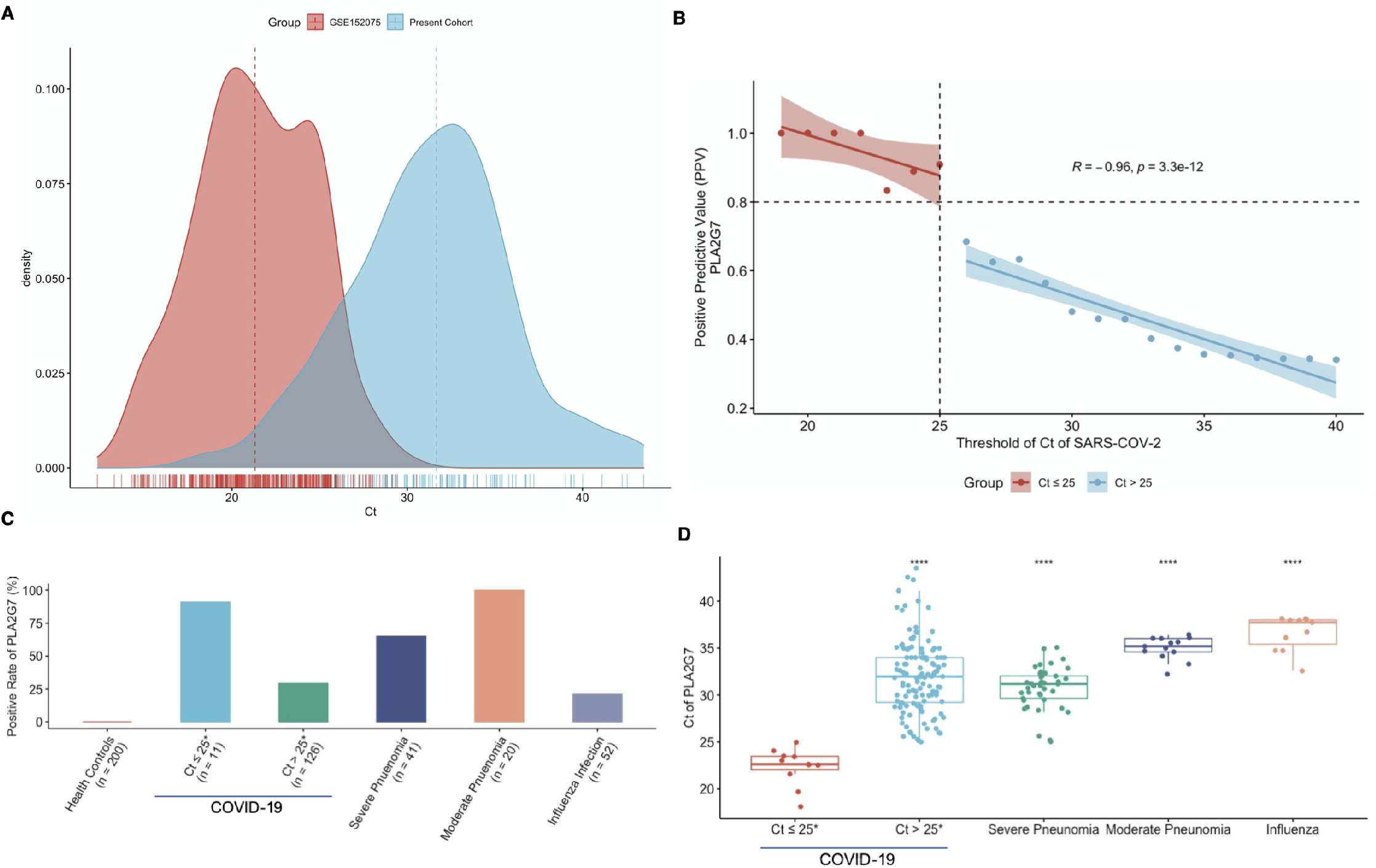
PLA2G7 was detected in a retrospective collection of nasal swabs from both COVID-19 and pneumonia patients. **(A)** Density plot of SARS-COV-2 Ct values distribution in GSE152075 and present cohorts. **(B)** Correlation between positive predictive value (PPV) of PLA2G7 and Ct values of SARS-COV-2. **(C)** Positive rates of PLA2G7 in six patients’ group. **(D)** Ct values distribution of PLA2G7 in patients’ group; The reference was COVID-19 group with Ct of SARS-COV-2 ≤ 25. ns: P ≥ 0.05; *: P < 0.05; **: P ≤ 0.01; ***: P ≤ 0.001; ****: P ≤ 0.0001.

The positive rates of PLA2G7 in all different groups were shown in Figure 3C. There were no PLA2G7 positive cases in health controls. Among the 134 SARS-COV-2 positive cases, 47 (35.1%) were PLA2G7 positive. It was worth of noting more than 90% PLA2G7 positive rate were observed in COVID-19 patients with Ct of SARS-COV-2 ≤ 25 while 29.37 % were seen in the group with Ct of SARS-COV-2 > 25 (Figure 3B-C). Surprisingly, a similar high detection rate of PLA2G7 was seen in patients with pneumonia (SRAS-CoV-2 negative). Notably, 100.00% of PLA2G7 positive rate were seen in group of severe pneumonia while 65.00% were noticed in moderate pneumonia patients. Conversely, only about 20.00% PLA2G7 positive rate were observed in patients with influenza infection. The Ct of PLA2G7 provided further insights (Figure 3D). The Ct values of PLA2G7 in group of SARS-COV-2 (Ct ≤ 25) were significantly lower than that in other groups. In addition, records in group with severe pneumonia showed higher levels of PLA2G7 than that in group with moderate pneumonia group (*P* = 7.2e-11) (Figure 3D; Supplementary Figure S12). It was worried to state that there were no significant differences between the Ct values of PLA2G7 in group of COVID-19 and that in severe pneumonia group (Figure 3D; Supplementary Figure S12).

### Abnormal serum protein levels of Lp-PLA_2_ in COVID-19 patients

PLA2G7 was also known as lipoprotein-associated phospholipase A_2_ (Lp-PLA_2_) in plasma(29). To understand the plasma levels of Lp-PLA2, a total of 117 serum samples (36 health controls, 81 COVID-19 patients) were collected. Among the 81 COVID-19 patients, 47 serum samples were collected at various time points from patients during the hospitalization (Hospitalization Group), 19 were from the patients who had been discharged and then re-positive but asymptomatic (Re-positive Group), and 15 were from fully recovered individuals (Recovered Group). Compared with the health controls, serum levels of Lp-PLA_2_ were higher in hospitalization and re-positive patients (Figure 4A). It was worth of noting that the hospitalization group showed significantly less levels of Lp-PLA_2_ than that in re-positive group (*P* = 0.004). Thereafter, we sought to explore the trend of Lp-PLA_2_ along with time. The overall trend of Lp-PLA_2_ in hospitalization group was reduced along with the days from initial symptoms onset (Figure 4B). To our surprise, higher plasma levels of Lp-PLA_2_ was in moderate than in severe patients (Figure 4B). In addition, the serum levels of Lp-PLA_2_ showed discriminating power between patients with COVID-19 and health individuals with AUC of 0.815 (95%CI 0.734 – 0.895) (Figure 4C).

**Figure 4.**
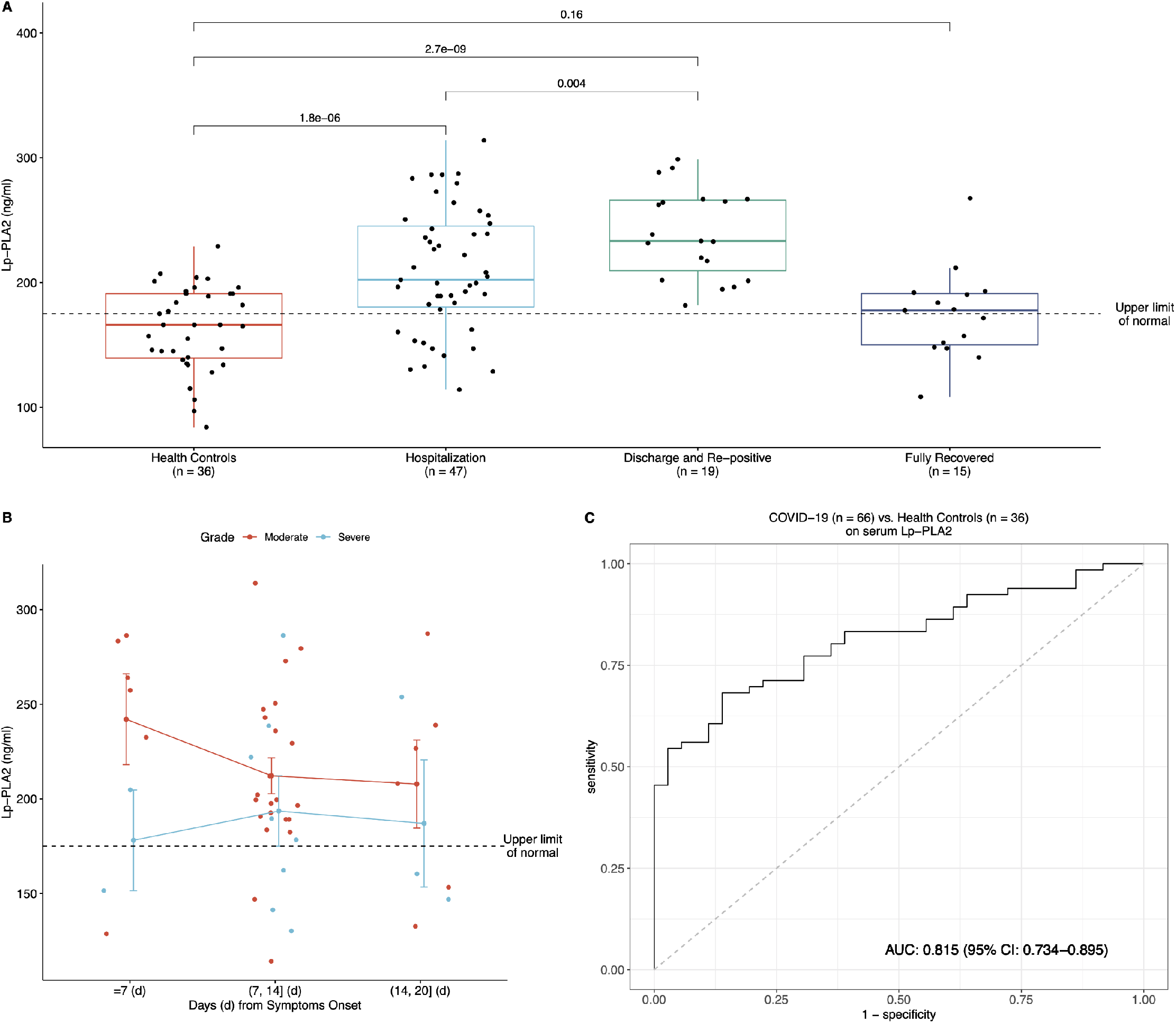
Abnormal plasma levels of Lp-PLA_2_ in COVID-19 patients. **(A)** Plasma levels of Lp-PLA_2_ in health controls (n = 36), hospitalized COVID-19 patients (n = 47), discharged and re-positive COVID-19 patients (n = 19) and fully recovered COVID-19 patients (n = 15). **(B)** Scatter plot of plasma levels of Lp-PLA_2_ in hospitalization group along with time from the initial symptoms onset. **(C)** Evaluation of classification performance of plasma levels of Lp-PLA_2_ between COVID-19 patients and health controls.

## Discussion

Given that the morbidity and mortality seen in COVID-19, a better understanding of the immunological underpinnings seen in patients infected with SARS-CoV-2 is necessary to better identify biomarkers, especially these represented the therapeutic and diagnostic targets. It has been suggested that cytokine blood RNA level was not always correlated with the protein plasma level(9). We therefore applied our previous established hypothesis-free single biomarker analysis framework on nasal swabs from more than 400 donors with COVID-19 in GSE152075. Finally, upregulated PLA2G7, a biomarker for cardiovascular diseases (CVDs)(29), was identified as a hub gene in SARS-COV-2 infection (Figure 1). Coincidentally, phospholipase A2 Group IID (PLA2G2D), belonged to the same PLA_2_ superfamily as PLA2G7 did, was considered as an increase factor to result in worse outcomes of mice infected by SARS-CoV(30). In addition, a 12-year follow-up survey of 25 patients who recovered from SARS-CoV infection found that 44% had cardiovascular system abnormalities(31) while almost half of hospitalized COVID-19 patients showed scan abnormalities that resemble the early stages of heart failure(12).

To understand the origin of PLA2G7, we re-analyzed scRNA-seq data from COVID-19 patients BALFs in GSE145926. We presented evidence-based findings that PLA2G7 was predominantly expressed by macrophages, especially the proinflammatory macrophages, including FCN1^hi^ group, FCN1^lo^SPP1^+^ group and SPP1^+^ group (Figure 2). These proinflammatory macrophages were emerging with progression of COVID-19 suggesting PLA2G7 could be used to monitor the progress of COVID-19 (Figure 2E-F). Previous studies suggested there was no mRNA of PLA2G7 in monocytes, but expression was induced and maintained during differentiation into macrophages(32). Therefore, our results might provide insights to the hypothesis that dysregulated activation of the mononuclear phagocyte (MNP) compartment contributes to COVID-19-associated hyperinflammation(33). Hence, the PLA2G7 which was specifically induced by proinflammatory monocyte-driven macrophages in lungs was seen in patients suffered from COVID-19.

We next validated the expression of PLA2G7 in more than 400 nasal samples by qRT-PCR. Comparing to the diagnostic performance of PLA2G7 in public dataset (Figure 1E), the PPV of PLA2G7 in SARS-COV-2 positive samples was relatively low. This discrepancy might be due to the relatively low viral load in present cohort (Figure 3A). It was noted that PPV of PLA2G7 in SARS-COV-2 positive samples were over 0.8 when the Ct of SARS-COV-2 was lower than 25 (Figure 3B). As proinflammatory macrophages in lungs played central roles in pneumonia(34), we postulated that PLA2G7 could be detected in not only patients with COVID-19, but also patients with pneumonia. The positive rate of PLA2G7 in nasal swabs from donors with pneumonia was higher than that in health controls and patients suffered seasonal influenza infection (Figure 3C). It was worth of noting that 100% positive rate of PLA2G7 were observed in severe pneumonia. In addition, the Ct values of PLA2G7 were significantly lower in severe pneumonia than in moderate pneumonia (p = 7.2e-11). This was consistent with our findings that expression of PLA2G7 was correlated with progression of COVID-19 (Figure 2F), suggesting PLA2G7 could be also used to monitor the progress of pneumonia. On-going epidemiological evidence substantiated the correlation between pneumonia and subsequent CVDs(35). Up to 30% of patients hospitalized for Community-acquired pneumonia (CAP) developed CVDs(36). Moreover, pneumonia has been suggested as a risk factor of CVDs(37). However, the patho-mechanisms by which pneumonia may trigger or promote subsequent CVDs still remain unclear. The expression profile of PLA2G7 was observed in patients with pneumonia could help to understand patho-mechanisms underlying pneumonia and CVDs. On the other hand, Ct values of PLA2G7 in group COVID-19 patients with high viral loads (SARS-COV-2 Ct ≤ 25) were significantly lower than that in the rest groups (Figure 3D). Importantly, no significant differences between the Ct values of PLA2G7 in group of COVID-19 patients with SARS-COV-2 Ct > 25 and that in severe pneumonia group. These data might provide an indication that more than half of COVID-19 hospitalized patients developed conditions with CVDs(12).

The abnormal levels of plasma protein of Lp-PLA_2_ have been seen in COVID-19 patients, especially the non-hospitalization patients (Figure 4). First of all, the aberrant plasma levels of Lp-PLA_2_ were observed in COVID-19 patients from not only hospitalization group, but also re-positive group (Figure 4A). Strikingly, the plasma level of Lp-PLA_2_ in hospitalization group was significantly lower than that in repositive group (p = 0.004) (Figure 4A). It was necessary to point out all the patients in re-positive group were asymptomatic. Therefore, the re-positive patients who had great potential risks on CVDs might, however, receive limited medical attention on CVDs. In addition, although there was no significant difference (p = 0.16) in the plasma level of Lp-PLA_2_ between fully recovered COVID-19 patients and health controls, it was worried to state that the mass of Lp-PLA_2_ in more than half (53.3%) of recovered patients (n = 15) was beyond the upper limit of normal (Figure 4A). Coincidentally, a recent study showed 78% of recovered COVID-19 patients, including illness ranging from asymptomatic to moderate symptoms, suffered issues association with the heart(38). Secondly, although the Lp-PLA_2_ was reduced along with time in hospitalized COVID-19 patients, it should be noted that the plasma levels of Lp-PLA_2_ in these patients were beyond the upper limit of normal, especially the moderate patients (Figure 4B). According to National Health Commission of China (NHC), some of the patients (later COVID-19 confirmed cases) first went to see a doctor because of cardiovascular symptoms, rather than respiratory symptoms(39). Even without symptoms and signs of interstitial pneumonia, cardiac involvement as a complication associated with COVID-19 has been reported^(40)^. Compare to severe group, the moderate patients who always presented fewer and/or lighter symptoms of CVDs leaded to limited medical care on CVDs. This might initiate follow-up comorbidities with heart in these patients.

To our knowledge, this is the first study that uncover the relationship between PLA2G7 and COVID-19 from both nucleotide and protein aspects. The results of our study provide important indications into the prevalence of cardiovascular involvements seen in COVID-19 patients, including status ranging from hospitalization to recovery period. In addition, our findings reveal the aberrant profile of PLA2G7 persists beyond the period of acute manifestation, especially in the discharged and re-positive patients. Importantly, the expression profile of PLA2G7 in COVID-19 patients with moderate pneumonia shows similar patterns seen in severe pneumonia, suggesting COVID-19 might enhance the expression of PLA2G7 (Figure 3). Although the long-term health effects of role of PLA2G7 in COVID-19 cannot yet be determined, on-going epidemiological evidence substantiated heart issues were occurred after COVID-19(41). Thus, our findings may provide an indication of potentially considerable burden of CVDs in large and growing parts of the population and urgently require confirmation in a larger cohort.

Nevertheless, our study has certain limitations. The expression of PLA2G7 was not validated in nasal swabs from the patients with severe COVID-19. In addition, the plasma levels of Lp-PLA_2_ in pneumonia patients were also not tested. Due to limited information of clinical traits of samples, confounding factors, such as pre-existing CVDs, which could increase the likelihood of PLA2G7 positivities, were not controlled. The in-house qRT-PCR assay of PLA2G7 was not optimized, leading to inadequate diagnostic performance. Most of the samples were retrospectively collected in Wuhan, China. It has been almost several months passed since all the samples were collected. The freshness of samples could also undermine the performance of the in-house PLA2G7 targeted qRT-PCR and commercial Lp-PLA_2_ ELISA toolkit.

Taken together, we firstly identified proinflammatory monocytes-driven macrophages specific PLA2G7 in lungs, a typically biomarker of CVDs, which leaded to abnormal levels of plasma Lp-PLA_2_, was enhanced in COVID-19. Importantly, PLA2G7 was detected in nasal swabs from patients with pneumonia. Although levels of PLA2G7 was significantly elevated in severe pneumonia comparing to moderate pneumonia, there were no significant differences between that in COVID-19 and severe pneumonia. Moreover, increased levels of Lp-PLA_2_ in plasma could provide insights to higher mortality was seen in patients underlying comorbidities (e.g. hypertension, diabetes mellitus, cardiovascular disease) and severe COVID-19 could occur in patients without pre-existing complications. Nevertheless, the role of PLA2G7 in the pathogenesis of COVID-19 worth cautious tests and studies.

## Materials and Methods

### Data collection

In brief, data were obtained from the Gene Expression Omnibus (GEO) database (http://www.ncbi.nlm.nih.gov/geo/) in July 2020 using the keyword “SARS-COV-2”. The following exclusion criteria were applied to the expression profiling by high throughput sequencing: (1) concerned only cell model; (2) no or insufficient clinical data; and (3) used non-baseline (“healthy”) controls. After review, GSE152075, which contained 484 samples of nasal swabs, was selected for biomarker discovery. Among them, in a total of 430 subjects were positive with SARS-COV-2 infections while 54 were negative. According to batch information, the batch effect hidden in the raw count matrix of GSE152075 was removing by function Combat_seq in the R package sva(42). Next, GSE145926 which included single-cell RNA sequencing (scRNA-Seq) data on bronchoalveolar lavage fluid (BALF) cells from three patients with moderate COVID-19, six patients with severe/critical infection and three healthy controls were also retrieved(27).

### Differentially expressed genes screening

When the batch-corrected count matrix of GSE152075 was compared in SARS-COV-2 positive cases with negative cases, DESeq2(19) was applied for differential expressed genes (DEGs). Thereafter, correction for multiple testing was addressed by controlling the false discovery rate (FDR) using the Benjamini and Hochberg (B.H.) method for both above. Criteria for DEGs were an absolute log_2_ fold change (Log_2_FC) of 0 and the FDR-adjusted *P*-value of < 0.05.

### Co-expression network construction

A co-expression network was constructed using the normalized GSE152075 data by the weighted correlation network analysis (WGCNA) in R (21). Briefly, quality assessment of GSE152075 samples was conducted using the cluster method. After removing outliners, the soft-thresholding power was then calculated, with the type of network set to signed. Network construction was then performed with correlation coefficient threshold was 0.85 (Default setting). In addition, the minimum number of genes in each module was 30 and the threshold for cut height was set to 0.25 to merge possible similar modules.

### Identification of modules related to SARS-COV-2 infection

For a given module, the expression profile was summarized into a single characteristic expression profile, designated module eigengenes (MEs). MEs were considered as the first principal component in the principal component analysis (PCA). Thereafter, a Pearson correlation analysis, calculating the student asymptotic *P-*values for the correlations, between MEs and clinical traits (SARS-COV-2 negative, SARS-COV-2 positive, Gender and Age) was conducted.

### Disease Ontology analyses

To understand the functions of enriched genes in interesting modules, Disease Ontology (DO)(22) analyses were performed using clusterProfiler(43), identifying significant results based on a *P*-value < 0.05 and gene counts ≥2.

### Candidate single gene biomarker selection

The module that was most highly correlated with SARS-COV-2 infection was selected. Candidate genes in the module were determined by both gene significance and module membership. Thereafter, a single gene biomarker was selected using XGBoost with recursive feature elimination with cross-validation (RFECV) (24, 25).

### Sample integration, dimensionality reduction and clustering on scRNA-data

Count matrixes from 10X CellRanger hdf5 in GSE145926 were obtained. The following criteria were then employed to filter low-quality cells: gene number between 200 and 6,000, unique molecular identifier (UMI) count > 1,000 and mitochondrial gene percentage < 0.1. Thereafter, the gene-cell matrixes of all samples were integrated with Seurat (version: 3) to remove batch effects across different donors. Next, the integrated gene-cell matrix was normalized using ‘LogNormalize’ methods in Seurat(44)with default parameters. The top 2,000 variable genes were then identified using the ‘vst’ method in Seurat FindVariableFeatures function. Variables ‘nCount_RNA’ and ‘percent.mito’ were regressed out in the scaling step. PCA was performed using the top 2,000 variable genes. Then UMAP (Uniform Manifold Approximation and Projection) and tSNE (t-distributed Stochastic Neighbor Embedding) were performed on the top 50 principal components (PCs) for visualizing the cells. Graph-based clustering was performed on the PCA-reduced data for clustering analysis with Seurat v.3. The resolution was set to 1.2 to obtain a finer result(27). Briefly, the first 50 PCs of the integrated gene-cell matrix were used to construct a shared nearest-neighbor graph (SNN; FindNeighbors() in Seurat) and this SNN was used to cluster the dataset (FindClusters()) using a graph-based modularity-optimization algorithm of the Louvain method for community detection.

### Macrophages re-integration

Macrophages of all samples were re-integrated using the first 50 dimensions of canonical correlation analysis and PCA with the parameter k.filter was set to 115. In the clustering step, parameter resolution was set to 0.8.

### Retrospective collection of clinical samples and case definition

We enrolled nasal swabs from donors with health condition and diagnosis of COVID-19, influenza infection and pneumonia. In a total of 447 nasal swabs were collected. Among these samples, 200 were from health individuals; 134 moderate pneumonia cases of confirmed diagnosis of COVID-19 (SARS-COV-2 positive) and 20 suspected COVID-19 cases (SARS-COV-2 negative) with moderate pneumonia were collected from government authorized hospitals for COVID-19 in Wuhan during the outbreak of COVID-19 (January 2020 to April 2020); 52 cases with H1N1 infection were also included in Wuhan from October to November, 2019 while 41 cases from hospitalized children who were diagnosed as severe pneumonia (SARS-Cov2 negative) were collected from Qilu Children’s Hospital, Jinan, Shandong in February, 2020. For the 117 serum samples, in a number of 81 serum samples from patients with COVID-19 and 36 from health donors were also collected from government authorized hospitals for COVID-19 in Wuhan during the outbreak of COVID-19. Among the 81 COVID-19 patients, 47 serum samples were collected at various time points from patients during the hospitalization, 19 were from the patients who had been discharged and then repositive but asymptomatic, and 15 were from fully recovered individuals. There were 34 moderate cases, 8 severe cases and 5 critical cases in the 47 serum samples from hospitalized patients. Disease severity of the COVID-19 patients were defined as moderate, severe and critical based on the Interim Guidance for Novel Coronavirus Pneumonia (Trial Implementation of Seven Edition) by the National Health Commission of China issued on 3 March 2020 (http://www.nhc.gov.cn/yzygj/s7653p/202003/46c9294a7dfe4cef80dc7f5912eb1989.shtml).

### Human studies

The research protocol was approved by the human bioethics committee of the Chinese Center for Disease Control and Prevention, and all participants provided written informed consent.

### qRT-PCR assay for SARS-COV-2 and PLA2G7

All the nasal swabs were sent to BSL-3 lab in Hubei CDC with the standard operational procedure (SOP) of the biosafety laboratory to re-test the SARS-COV-2 RNA before detection of PLA2G7. Total nucleic acid was extracted using the QIAamp viral RNA mini kit (Qiagen) and SARS-COV-2 targeted qRT-PCR was performed using a China Food and Drug Administration-approved commercial kit specific for SARS-CoV-2 detection (Da An Gene). This kit is based on one-step TaqMan qRT-PCR technique, and ORF1ab and N genes are selected as amplification target regions. Briefly, the 25-µL reaction mixture comprised 17 µL of PCR reaction Solution A, 3 µL of PCR reaction Solution B, and 5 µL of RNA extracts. The cycling conditions were as follows: 50 °C for 15 min, 95 °C for 15 min, followed by 45 cycles for 15 s at 94 °C and 45 s at 55 °C. Each qRT–PCR assay provided a cycle threshold (Ct) value, indicating the number of cycles surpassing the threshold for a positive test. The Ct value for a positive specimen was set at 40 cycles.

To study PLA2G7 transcripts expression, the primers and TaqMan probes selected from the Universal ProbeLibrary Assay Design Center (Roche Diagnostics) were applied. In briefly, the TaqMan qRT-PCR was carried out in AgPath-ID™ One-Step RT-PCR Reagents (Applied Biosystems) using a CFX96^TM^ Real-Time PCR Detection System (Bio-Rad). Cycling conditions were 45°C for 10 min and 95°C for 10 min, followed by 40 cycles of 95°C for 15 s, 58°C for 45 s. At least three replicates were studied for each sample. The PLA2G7 expression levels were normalized to RHOG, and the relative expression was calculated as 2^−ΔΔCt^.

### Measurement of Plasma levels of Lp-PLA2

The plasma levels of Lp-PLA_2_ were detected by enzyme linked immunosorbent assay (ELISA) according to the instruction (Hotgen, registration no. 20192400366). In brief, serum was heat-inactivated at 56℃ for 45 min. Each sample (20 µL) and standard were added to a 96-well plate. A total of 100 μl of enzyme-labeled reagent was added to all wells (except the blank well) and incubated at 37 °C for 1 h. The plate was washed 5 times before chromogenic reagents A and B (50 μl of each) were mixed and added to each well. The plate was incubated in the dark at 37°C for 10 min before 50 μl stop solution was added to each well to terminate the color reaction. The optical density (OD) 450 nm values in each well were determined by a microplate reader. The levels of Lp-PLA_2_ were then calculated according to the standard curve. The upper limit of normal level of Lp-PLA_2_ were 175 ng/ml.

### Statistical analysis

R (version 4.0.2) was used for most analyses, with hub gene selection being performed using XGBoost and Scikit in Python (version 3.6). Differences of median percentage among in group were compared using a Student’s t-test (one-sided [less], unadjusted for multiple comparisons) with R ggpubr v.0.2.5. P-value ≤ 0.05 was considered statistically significant.

## Data Availability

The datasets GSE152075 and GSE145926 for this study can be found in the Gene Expression Omnibus (GEO) database hosted by the National Center for Biotechnology Information of the US National Institutes of Health (https://www.ncbi.nlm.nih.gov/geo/). All data are available in the main text or the supplementary materials.

## Author Contributions

SW, ZG, FZ and ML conceived and designed the study and had full access to all of the data in the study. Yang L, YJ, YZ, NL, SW, ML drafted the paper. Yang L, YJ, YZ, NL, QY, Yan L, AL, JL, XC, JL, DL did the analysis. LL, XL, BF, HY, GY banked the samples. QY, Yang LIU, YQ, CL performed experiments. All authors critically revised the manuscript for important intellectual content and gave final approve for the version to be published. All authors agree to be accountable for all aspects of the work in ensuring that questions related to the accuracy or integrity of any part of the work.

## Acknowledgments

We acknowledge all health-care workers involved in the diagnosis and treatment of patients in Wuhan and Jinan. We are grateful for all the patients for their supports and confidence in our study. We thank the researchers who obtained the sequencing data used in this study. We thank the helpful discussions with Xianbing YU from the Chemistry Department at the University of Chicago and Chen CHEN from Beijing Ditan Hospital, Capital Medical University. We thank Yiming ZHOU and Yunke LI from Beijing Neoantigen Biotechnology Co. Ltd for their helps in single cell data analysis. We thank the valuable comments about PLA2G7 from clinical doctors, especially Guiqiang WANG from Peking University First Hospital and Jun ZHU from Fuwai Hospital, Chinese Academy of Medical Sciences.

